# SMELL-RS: A Self-administered, Digital Test for Olfactory Dysfunction that is Rapid, Reliable, and Accurate

**DOI:** 10.64898/2026.03.28.26349316

**Authors:** Julien W. Hsieh, M Dougherty, Aikaterini Poulopoulou, Déborah Blidariu, Pascal Senn, Richard Hopper, Devarsh Patel, Emanuela Maggioni, Marianna Obrist, Leslie B. Vosshall, Andreas Keller, Basile N. Landis

## Abstract

**Background:** Smell testing is increasingly recognized as essential in rhinology practice but remains underutilized because of time constraints and limited clinical resources. This study aimed to evaluate the performance (test-retest reliability, accuracy and test completion time) of a self-administered, digital version of SMELL-RS, a non-semantic test of olfactory resolution (SMELL-R) and sensitivity (SMELL-S).

**Methodology:** We performed a test-retest reliability study in a tertiary care facility. We enrolled 100 subjects with and without smell dysfunction. The primary outcome measures were two replicates of olfactory test scores (SMELL-RS composite score, SMELL-R score, SMELL-S score). The secondary outcome measures were Sniffin’ Sticks score, test completion time, patient demographics, and other clinical characteristics (clinical symptoms, etiologies).

**Results:** The SMELL-RS composite score was reliable (ICC=0.71; p<0.0001) and correlated with the Sniffin’ Sticks composite score (r=0.68; p<0.0001). Different etiologies have different magnitudes of smell loss as revealed by the SMELL-RS score. SMELL-S reduces misdiagnosis associated with Sniffin’ Sticks threshold tests. The average completion time of the olfactory resolution test (SMELL-R) was on average 5.9 minutes (SD=1.9), while the average completion time of the olfactory sensitivity test (SMELL-S) was 5.5 minutes (SD=2.7). This is two to three times faster than the corresponding Sniffin’ Sticks tests.

**Conclusions:** SMELL-RS is a rapid, fully automated, reliable, and accurate olfactory test suitable for self-administration in a clinical setting.

## INTRODUCTION

Over a century of innovation in olfactory testing has yielded more than one hundred tests, yet no international consensus exists on their use (1, 2). Two major barriers explain this: test inaccuracy linked to cultural or genetic differences, and the limited practicality of current smell tests (3-5). They limit global collaborative science and the adoption of smell tests in clinical practice.

Regarding the first barrier, olfactory threshold tests, analogous to pure-tone audiograms, measure the lowest detectable concentration of a mono-molecular odorant, such as phenylethyl alcohol that smells like rose. Unlike in hearing, the sensitivity to specific olfactory stimuli varies greatly among healthy individuals (6, 7). This is partially due to genetic variability in specific odorant receptor genes (7, 8). This variability in the healthy population limits the interpretation of olfactory threshold test scores using mono-molecular odorants (9). Other methods analyse the ability to match an odor to its correct label (identification test) or to tell apart different odors (discrimination test). They are biased by pre-existing familiarity with both odor and specific label being tested, making them poorly adaptable across cultures (10-12). These genetic and cultural biases limit universal adoption of existing tests.

Regarding the second barrier, widely used tools such as the T&T Olfactometer (Japan), UPSIT (North America), and Sniffin’ Sticks (Europe), which use at least one or a combination of the tests mentioned above, have advanced the field through standardization (13-15). They are mostly employed in tertiary hospitals with specialized smell and taste clinics. Their administration takes up to 45 minutes per patient and requires trained staff to administer the tests. Because of this, these tools are absent from most clinical settings, and most hospitals and private practices instead use rapid, low-resolution screening tests that can distinguish normal from impaired olfaction (16). These types of screening tests are rapid but lack the granularity and accuracy needed for optimal clinical management. Despite the superior accuracy of tests that take 45 minutes to complete, only 17% of surveyed otorhinolaryngologists would consider performing a 15–45 minute test in a perioperative setting (3).

To address the limitations due to olfactory bias, we utilized “olfactory whites”, which are complex odor mixtures of over 20 equi-intense odorants spanning the physico-chemical odor space (17) (Figure 1a). They may activate a broader range of odorant receptors and typically have unfamiliar smells. This led to the creation of a universal testing method called SMELL-S (sensitivity) and SMELL-R (resolution) tests (Figure 1b,c), which were shown to be reliable, accurate, and universal (9). The first prototype used a self-administered system with a software app, tablet, barcode scanner, and a large tray containing many barcoded glass vials. This prototype was not practical for clinical use. Therefore, we describe an approach with a recently developed multi-channel portable odor delivery device for self-administered and rapid smell testing in a clinical setting. This device can deliver up to 24 different odor stimuli with high temporal precision and without cross-contamination. It allows for the rapid sequencing of odors via digital control using a computer (Figure 1d) (18). Such technology may facilitate the adoption of diagnostic smell testing in different clinical settings.

**Figure 1.**
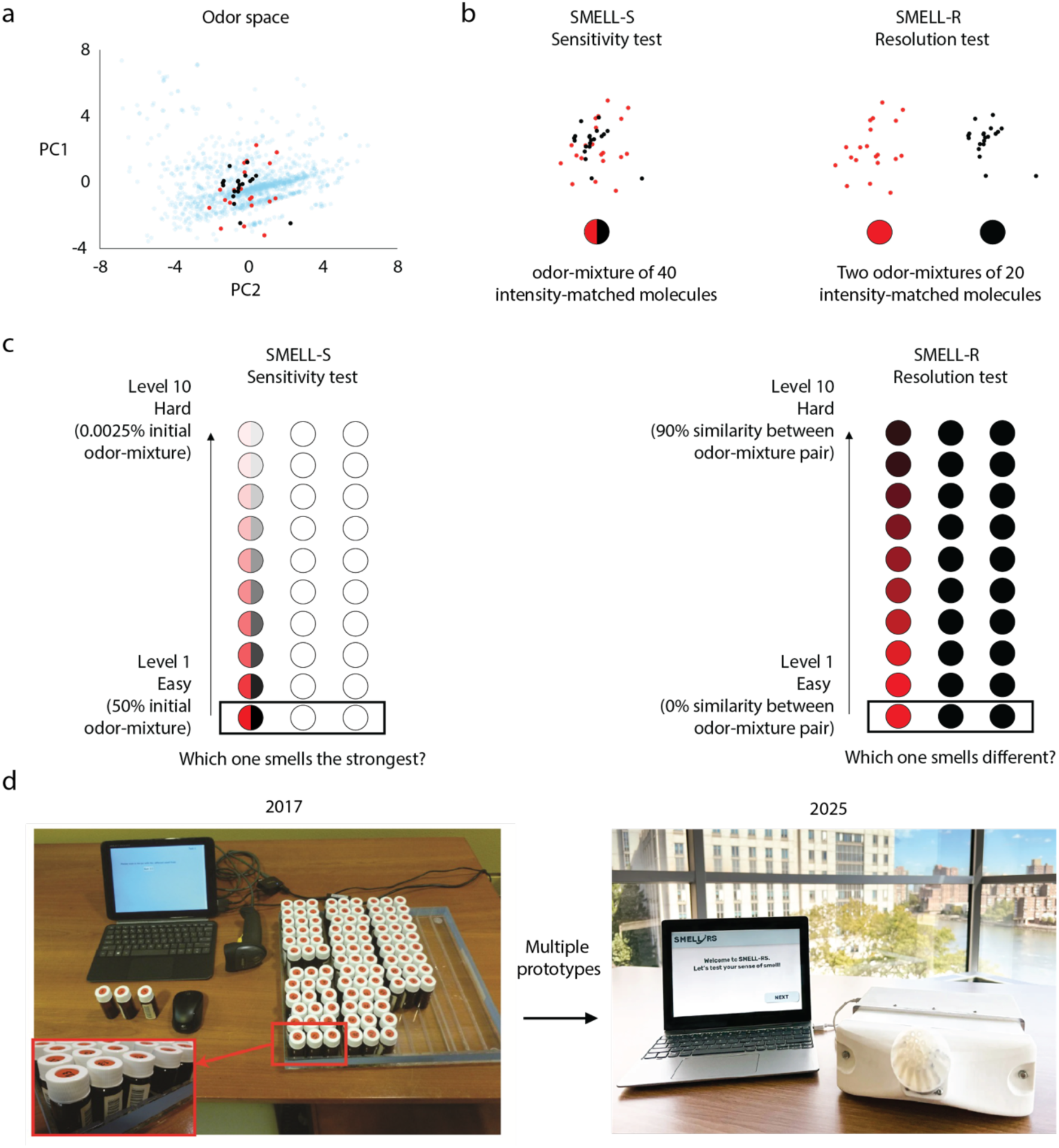
SMELL-RS concept. (**a**) Odor space representing thousands of molecules (blue dots), for which 1664 physicochemical descriptors were calculated and projected onto a 2-dimensional space made of first and second principal components (PC). The more distant two molecules are, the more physico-chemically different they are. 40 molecules were selected and intensity-matched (black and red dots). (**b**) The molecules used in the SMELL-S stimulus (left) and the molecules used for the two SMELL-R stimuli at level 1 (right). They were isolated from the 2-dimensional space made of two PCs of figure 1a. (**c**) Test design using a triangle test and an adaptive staircase threshold to determine the limit of perception. Both tests have 10 levels of difficulty that differ in levels of serial dilution (SMELL-S) or % of shared molecules between odor pairs (SMELL-R). (**d**) The barcoded vials used in the original version of SMELL-RS (left) and the digital olfactometer used for this study (right).

This study aimed to evaluate the test completion time, test–retest reliability, and diagnostic accuracy of the self-administered, digital version of SMELL-RS, in comparison with the Sniffin’ Sticks test.

## MATERIALS AND METHODS

### Experimental design

This prospective test-retest reliability study was performed between February 2023 and July 2024 in a tertiary care facility at the Rhinology-Olfactology Unit, Department of Otorhinolaryngology, of Geneva University Hospitals. The study included two visits spaced 1 week ± 3 days apart. At the first visit, the informed consent form was discussed and signed. Then, patients underwent a REDCap questionnaire focused on history taking, nasal endoscopy, SMELL-S, SMELL-R, and the Sniffin’ Sticks test (threshold, discrimination, identification). The Sniffin’ Sticks test was not performed if the patient previously had this test within 10 days in the context of their clinical management. At the second visit, the questionnaire and all the tests were repeated except the Sniffin’ Sticks discrimination and threshold tests. The order of the tests was randomized (randomizer.org). The study was approved by the institutional ethics review board and was conducted according to the Declaration of Helsinki on Biomedical Research Involving Human Subjects (IRB approval No: 2020-02581).

### Study subjects

We recruited 100 subjects aged 18 years and older to enroll. Subjects with normosmia were defined as having a Sniffin’ Sticks TDI composite score equal to or above 30.5 points and subjects with smell dysfunction had a Sniffin’ Sticks TDI composite score below 30.5. Patients were excluded if they were not able to understand instructions given by the investigators. They were screened by a consultation at The Rhinology-Olfactology Unit at the Geneva University Hospitals (n=57) or recruited through hospital website advertisements (n=43). All the visits and data management were carried out by co-authors (A.P, D.B) and HUG NeuroCentre, which supports clinical research in the hospital.

#### Outcome measures and procedures

### Patient characteristics

A REDCap questionnaire was created to record a subject’s history including detailed age, gender, ethnicity, geographic/cultural background, chemosensory and rhinological symptoms. This French questionnaire can be obtained upon request from the corresponding author. The most likely diagnosis was based on recent guidelines (2).

### Sniffin’ Sticks

Patients were tested using Sniffin’ Sticks (Burghart, Wedel, Germany), which includes olfactory threshold (T), discrimination (D), and identification (I). The TDI composite score differentiates anosmia, hyposmia, and normosmia. When each nostril was tested separately, the higher of the two TDI composite scores was used, according to established procedures (15, 19-21). The test completion time was manually recorded with a chronometer.

### SMELL-RS Software

We created software that allows self-administration of a series of triangle tests. A triangle test consists of two identical stimuli and one odd stimulus presented in randomized order. The subjects were instructed to either select the odor stimulus that was strongest (SMELL-S) or different from the others (SMELL-R). The stimuli each lasted 3 seconds, separated by a 3-second inter-stimulus interval. Then, whenever the subjects felt ready, they pressed “next” to start the next triangle test. The test uses an adaptive staircase paradigm identical to the Sniffin’ Sticks threshold test with seven reversals. A reversal is the trial where the stimulus progression changes direction (from becoming easier to harder or vice versa), indicating that the procedure may have crossed the participant’s threshold region. The software recorded the time (minutes) for each triangle test and the subject’s response.

### Digital Smell delivery device

The digital smell delivery device is a portable and affordable system designed to deliver up to 24 odors through independent channels with millisecond-level temporal precision (18). Each odor is stored in a sponge-based reservoir housed within a removable cartridge. The design minimizes contamination by using individual Teflon outlet pipes for each odorant, reinforced with rubber O-rings to ensure pneumatic sealing. The small headspace in each reservoir allows for rapid saturation of vapors, enabling quick and reliable odor delivery upon airflow activation. This multi-channel setup allows researchers to sequence odors digitally with high flexibility and to incorporate odorants of different types and concentrations without risk of cross-contamination (Figure 1d).

### SMELL-S – Sensitivity test

Analogous to an audiogram, which measures the minimum intensity at which a pure tone becomes perceptible, the olfactory threshold refers to the lowest concentration of an odorant that can be perceived. The stimulus used for SMELL-S is a mixture of 40 iso-intense odor molecules spanning the physicochemical space (Figure 1b). Note that the specific odorants used in this study differ from those in Hsieh et al. (2017) and can be obtained on request from the corresponding author (9). We created 10 levels of difficulty with 10 serial dilutions (Figure 1c). To know whether the stimulus can be perceived at a given level, a triangle task is used to deliver two odorless solvent stimuli and the odd odor in a randomized order. After each task, patients determine which odor smells the strongest. Olfactory threshold is reached by using an adaptive staircase threshold paradigm with seven reversals. Based on the collected data on test completion time, reliability, and accuracy, the SMELL-S score is defined as the mean of the last two reversals out of five total reversals (Supplemental file 1).

### SMELL-R – Resolution test

Analogous to visual resolution, which describes the minimum distance at which two points of light can be distinguished, smell resolution refers to the perceptual threshold at which two chemically similar odor mixtures can be discriminated as distinct rather than perceived as the same. The odor stimuli are composed of two mixtures of 20 odor molecules each spanning the physicochemical space (Figure 1b). Note that the specific odorants used in this study differ from those in Hsieh et al. (2017) and can be obtained on request from the corresponding author (9). We also created 10 odor-mixture pairs of increasing difficulty as the proportion of shared odor molecules increases (Figure 1c). To determine whether the odor pair can be distinguished at a given level, a triangle task is used to deliver two identical odor stimuli and one that is different in a randomized order. After each task, patients determine which odor smells different. The resolution is reached by using an adaptive staircase threshold paradigm with seven reversals. Based on the data collected on test completion time, reliability, and accuracy, the olfactory resolution score is defined as the mean of the last two (instead of four compared to Sniffin’ Sticks) of four staircase reversals (Supplemental file 1).

### SMELL-RS Composite score

The composite score is the sum of SMELL-R and SMELL-S scores.

### Statistical Analysis

Demographic, clinical, and psychophysical variables were compared between normosmic participants (n = 51) and those with smell dysfunction (n = 49). Continuous variables with normal distributions (e.g., age, Sniffin’ Sticks scores, SMELL-RS, SMELL-S) were analysed using unpaired t-tests, while non-normally distributed variables (e.g., nasal symptom scores, SMELL-R) were compared using the Mann–Whitney U test or Wilcoxon signed-rank test when appropriate. Categorical variables (gender, ethnicity, cultural identification, clinical complaints, and most likely diagnosis) were compared using the χ² test. Test–retest reliability was evaluated using the intraclass correlation coefficient (ICC) based on scores obtained at visit 1 and visit 2. Simple linear regression was also used to study these associations. Diagnostic accuracy was assessed using Spearman correlation, receiver operating characteristic (ROC) analyses, calculation of sensitivity, specificity, and the Youden Index. Differences in smell testing scores across etiological groups of olfactory dysfunction were analysed using the Kruskal–Wallis test. Test completion time was summarised using descriptive statistics. A p-value < 0.05 was considered statistically significant. We performed all analyses with GraphPad Prism 10.4.0. Adobe Illustrator 2025 was used to refine the figures.

## RESULTS

### Demographics and clinical characteristics

The characteristics of the study subjects are depicted in Table 1. The normosmic group included 33% of patients with post-infectious olfactory loss who had recovered.

**Table 1.**
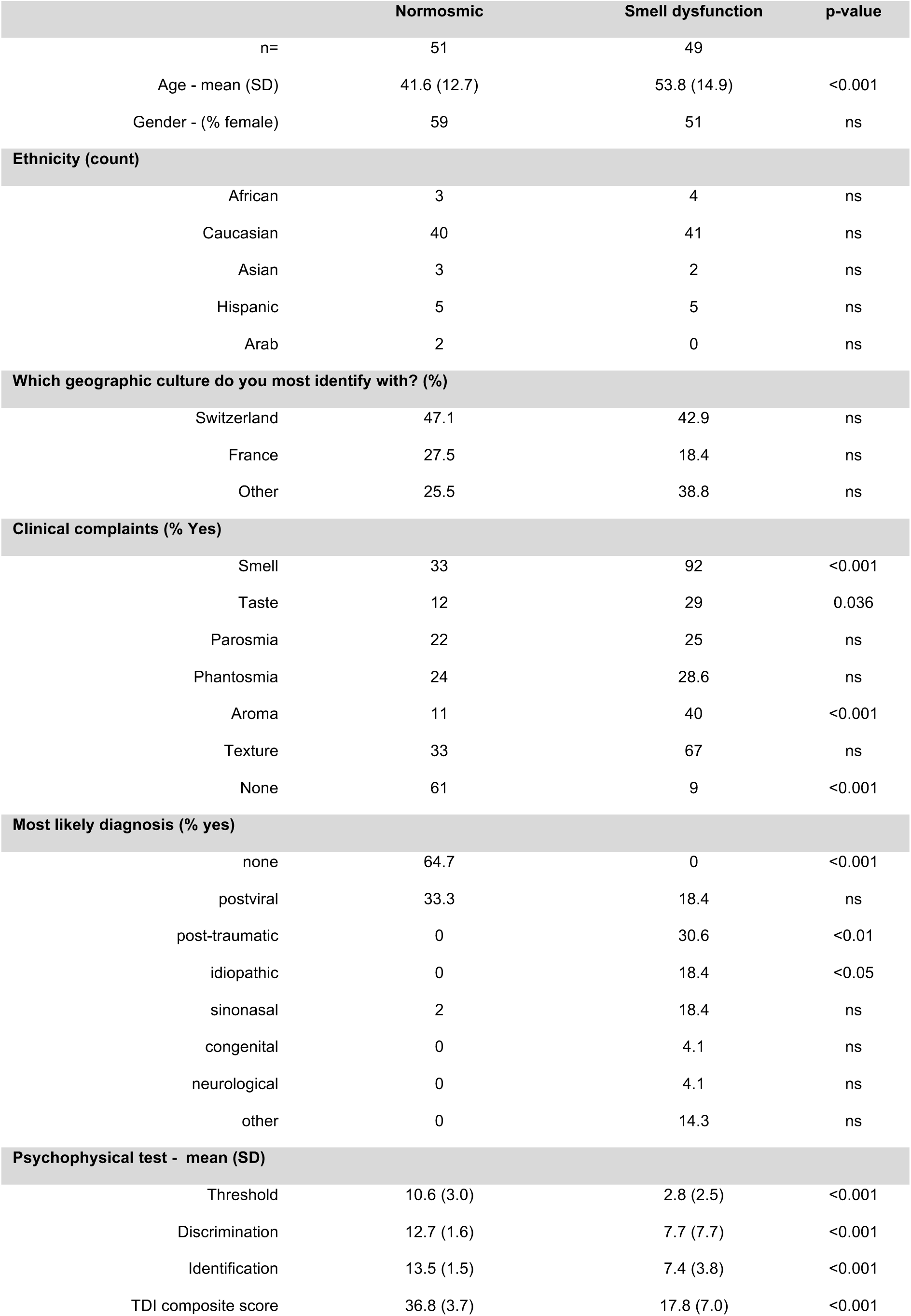
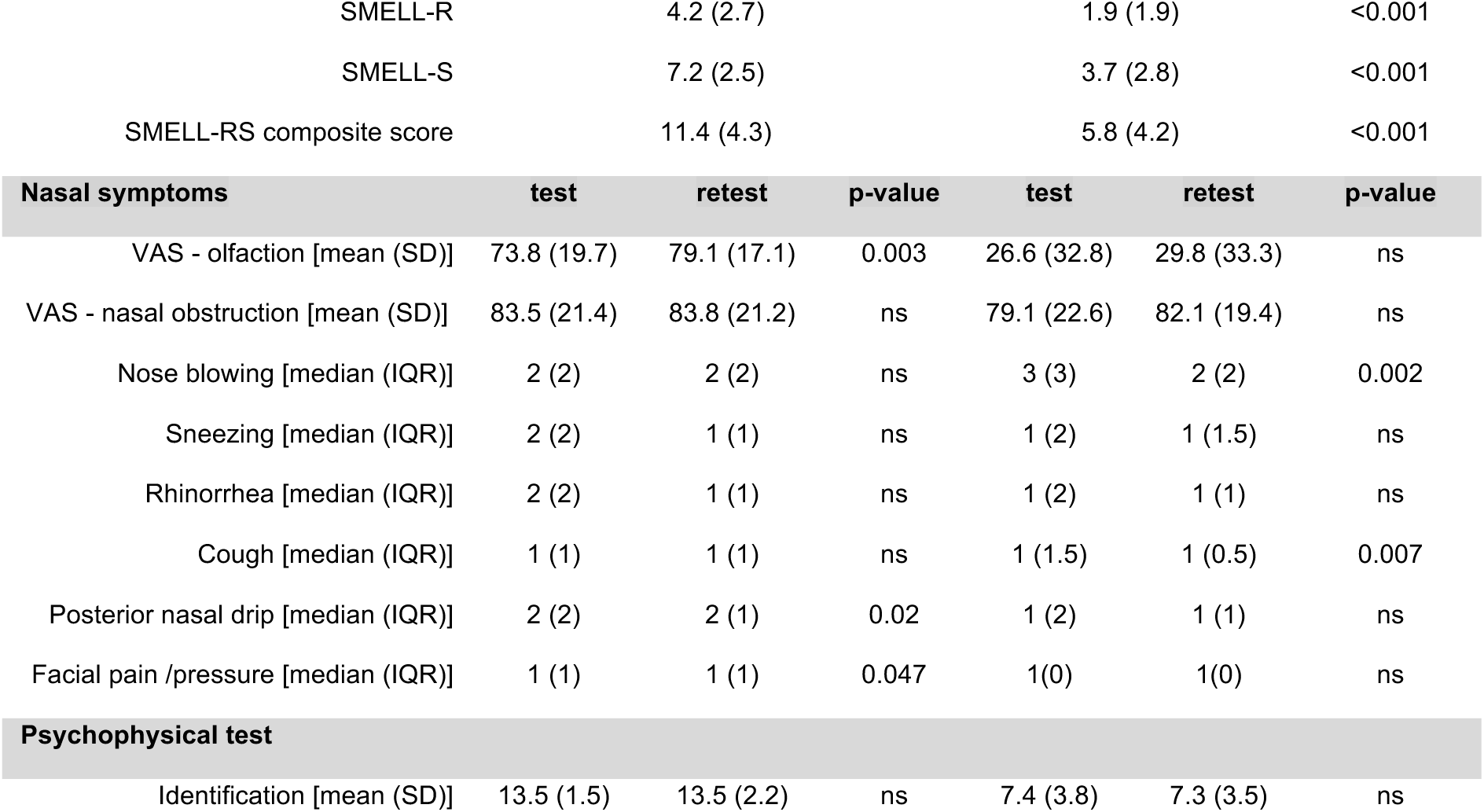
Demographics and clinical characteristics. The cultural group “other” includes: Algeria (n=2), Bahamas (n=1), Belgium (n=1), Bolivia (n=1), Brazil (n=1), Chili (n=2), Egypt (n=1), Spain (n=1), Greece (n=3), India (n=1), Italy (n=7), Laos (n=1) Morocco (n=1), Philippines (n=1), Portugal (n=3), Romania (n=3). ns=non-significant. SD=standard deviation. IQR=Interquartile range.

### Test-retest reliability and accuracy

We found that the Intraclass Correlation Coefficient (ICC) between test and retest scores for SMELL-S and SMELL-R were moderate, whereas it was strong for SMELL-RS composite score (Figure 2a). In addition, the correlations between the Sniffin’ Sticks TDI composite score and the SMELL-RS composite score, SMELL-R, and SMELL-S were moderate (Figure 2b). To identify optimal cut-off values for distinguishing individuals with normal olfaction from those with olfactory dysfunction, we calculated sensitivity, specificity, and the Youden Index. For the SMELL-RS composite score, we selected a cut-off of 7.6, which maximized both sensitivity (77%) and specificity (80%) (Youden Index = 0.57; AUC = 0.84, 95% CI = 0.76–0.92). After determining overall olfactory function using the composite score, the administering clinician can examine specific olfactory domains. For SMELL-R, which assesses olfactory resolution, we chose a cut-off of 3.75 to maximize sensitivity (89%) and facilitate ruling out olfactory dysfunction when scores exceed this threshold (specificity = 54%; Youden Index = 0.43; AUC = 0.79, 95% CI = 0.69–0.88). For SMELL-S, which measures general olfactory sensitivity, we selected a cut-off of 4.25 to maximize specificity (86%) and improve the ability to rule in dysfunction when scores fall below this value (sensitivity = 58%; Youden Index = 0.44; AUC = 0.82, 95% CI = 0.74–0.90) (Figure 2c).

**Figure 2.**
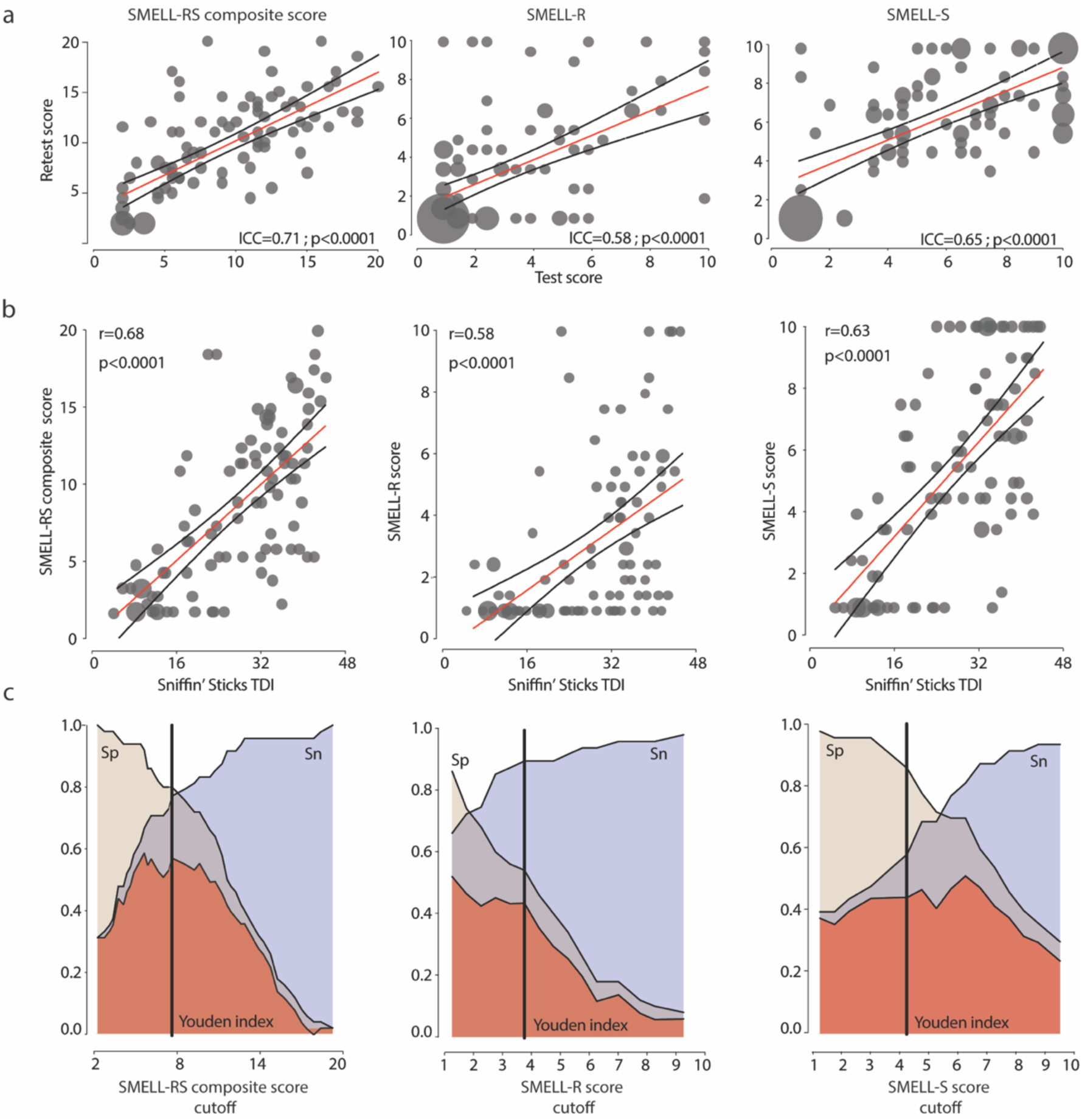
SMELL-RS is reliable and accurate. (**a**) Test-retest reliability. (**b**) Correlation between SMELL-RS and Sniffin’ Sticks. Each dot represents a study subject. The size of the dots are proportional to the number of subjects with the same overlapping score. ICC=Intraclass Correlation Coefficient. r=Spearman r. A simple linear regression line (red line) with 95% Confidence interval (black line) is depicted. (**c**) Test accuracy and cutoff determination. The vertical black line represents the chosen cut-off. Sp=Specificity, Sn=Sensitivity.

### SMELL-RS Differentiates Etiologies of Olfactory Loss

Because diagnosing the cause of olfactory impairment can be challenging, we grouped patients according to their most likely etiology and examined their SMELL-RS composite scores, as well as their SMELL-R and SMELL-S subtest scores. We found that the magnitude of smell loss, reflected by the SMELL-RS composite score or by SMELL-S alone, differed significantly between patients with post-infectious, head-trauma–related, and idiopathic olfactory dysfunction. This indicates that the magnitude of impairment may aid clinical diagnosis (Figure 3a). In clinical practice, we propose first evaluating the SMELL-RS composite score to determine whether a patient is normosmic, hyposmic, or anosmic. Subsequently, the SMELL-R and SMELL-S subtest scores can be interpreted separately to assess specific domains of olfactory function - resolution and general sensitivity, respectively (Figure 3b).

**Figure 3.**
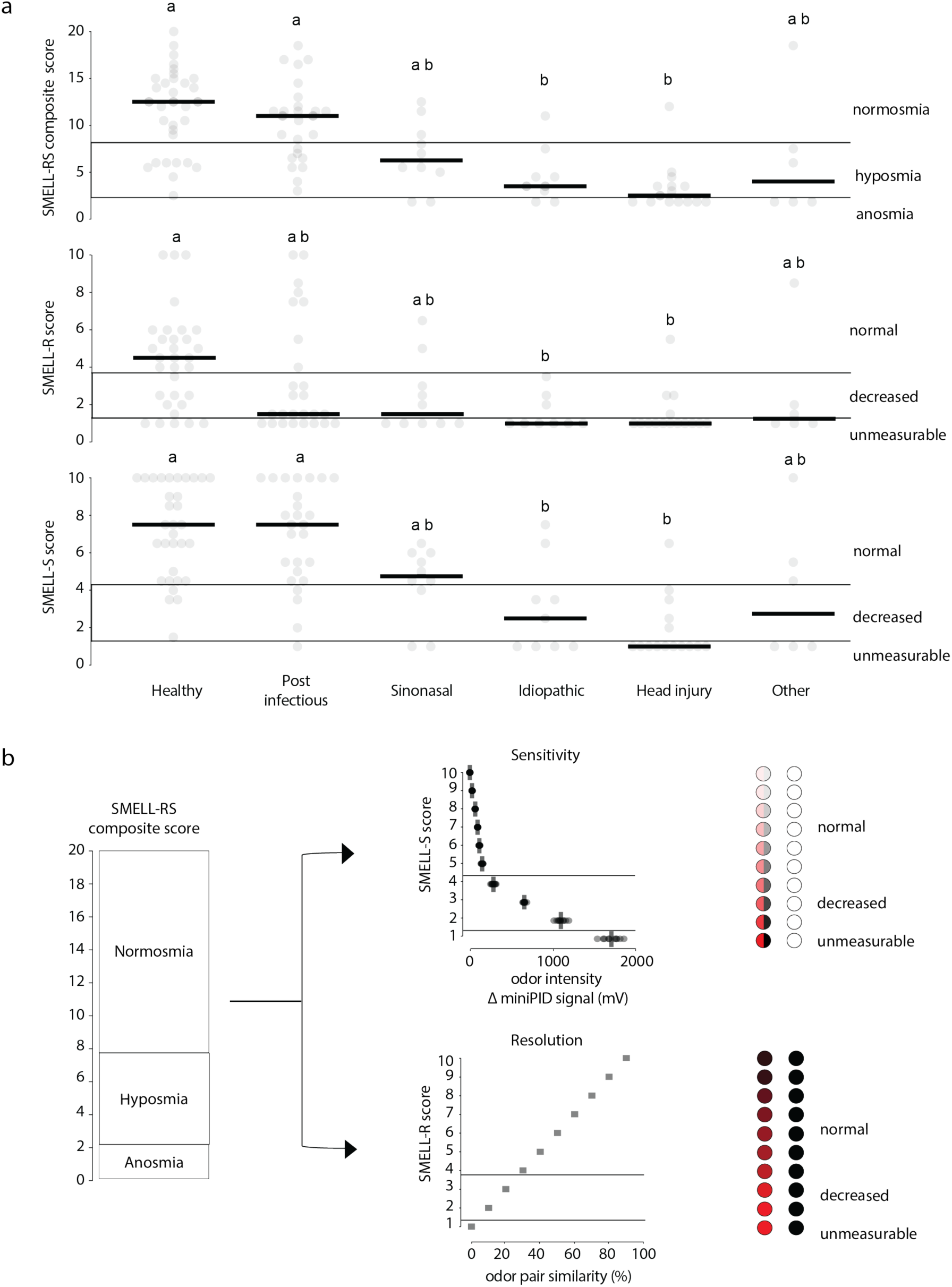
Clinical utility beyond quantifying smell loss. (**a**) Magnitude of olfactory deficit, as measured by SMELL-S and SMELL-RS composite score but not SMELL-R, is associated with specific putative etiologies of smell loss. Each dot represents a study subject. The median is depicted by a black bar. Groups labeled with different letters are significantly different (p≤0.05). (**b**) Algorithm for clinical usage and correlation with physical scale.

### SMELL-S reduces misdiagnosis associated with a Sniffin’ Sticks threshold test

Sensitivity to a single odorant, such as phenylethyl alcohol (a rose-like odor), can be inherently low due to natural genetic variation in odorant receptor genes across the population—variability that is unrelated to disease. To assess whether this limitation could lead to misdiagnosis in a clinical setting, we examined individuals who scored at the lowest level on the phenylethyl alcohol Sniffin’ Sticks threshold test (n = 25), a result that would typically suggest functional anosmia. We found that 44% (11/25) of these individuals showed moderately decreased olfactory function, or even normal function, when their threshold was measured using a 40-component odor-mixture (Figure 4a). Consistently, this group also reported higher self-rated olfactory ability and had better discrimination and identification scores (Figure 4b). Together, these findings indicate that clinically-relevant odor sensitivity is more accurately assessed using odor mixtures rather than single molecules. SMELL-S is therefore expected to lead to fewer false positives than the current clinical standard of the phenylethyl alcohol Sniffin’ Sticks threshold test.

**Figure 4.**
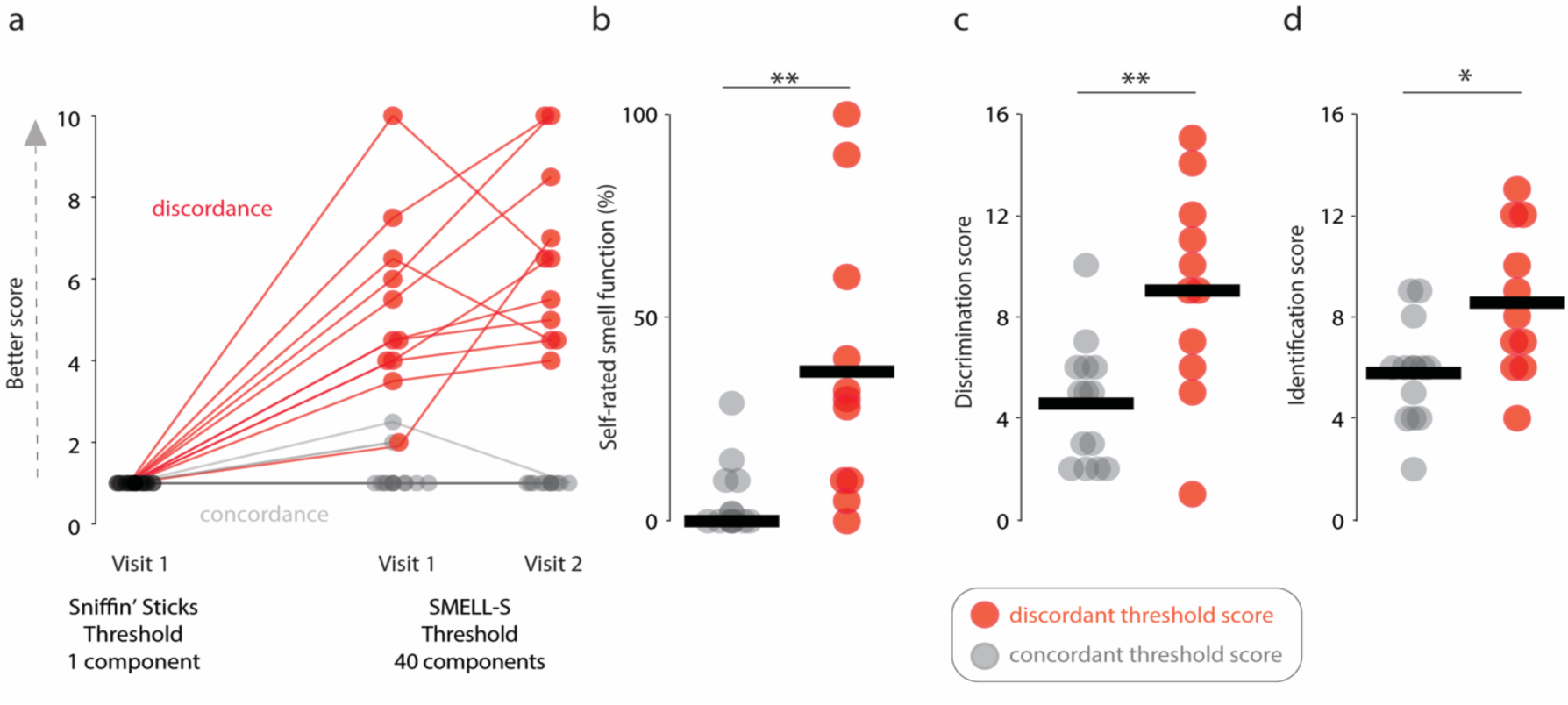
SMELL-S overcomes misdiagnosis encountered with the Sniffin’ Sticks phenylethyl alcohol threshold test. (**a**) All study subjects tested with the Sniffin’ Sticks phenylethyl alcohol threshold score are represented by a black dot (n=25). Their performance with the SMELL-S at test and retest visits is shown. A total of 11 out of 25 patients depicted with red dots and lines had a measurable SMELL-S threshold (discordant). These 11 patients had one of the following diagnosis: head trauma, post-infectious, idiopathic, sinonasal, other. 14 patients depicted in grey dots and lines had no measurable SMELL-S threshold (concordant). (**b-d**) Performance on other markers of olfactory function in patients with concordant or discordant threshold score.

### Test completion time

To achieve universal smell assessment, test completion time must be minimized to increase adoption rate in clinical settings but without sacrificing precision - a balance that is often difficult to achieve. Data were collected for n=79 subjects in the Sniffin’ Sticks composite score group, n=76 in the threshold group, n=78 in the discrimination group, n=79 in the identification group, n=98 in the SMELL-RS composite score group; n=100 in the SMELL-R group and n=98 in the SMELL-S group. The average completion time of the olfactory resolution test (SMELL-R) was 5.9 minutes (SD=1.9), while the average completion time of the olfactory sensitivity test (SMELL-S) was 5.5 minutes (SD=2.7) (Figure 5). Our data further suggest that for SMELL-S, the completion time can be even less than 5.5 minutes by lowering the number of reversals, with minimal impact on reliability and accuracy (Supplemental file 1). This is less than half the time needed to complete the corresponding Sniffin’ Sticks tests.

**Figure 5.**
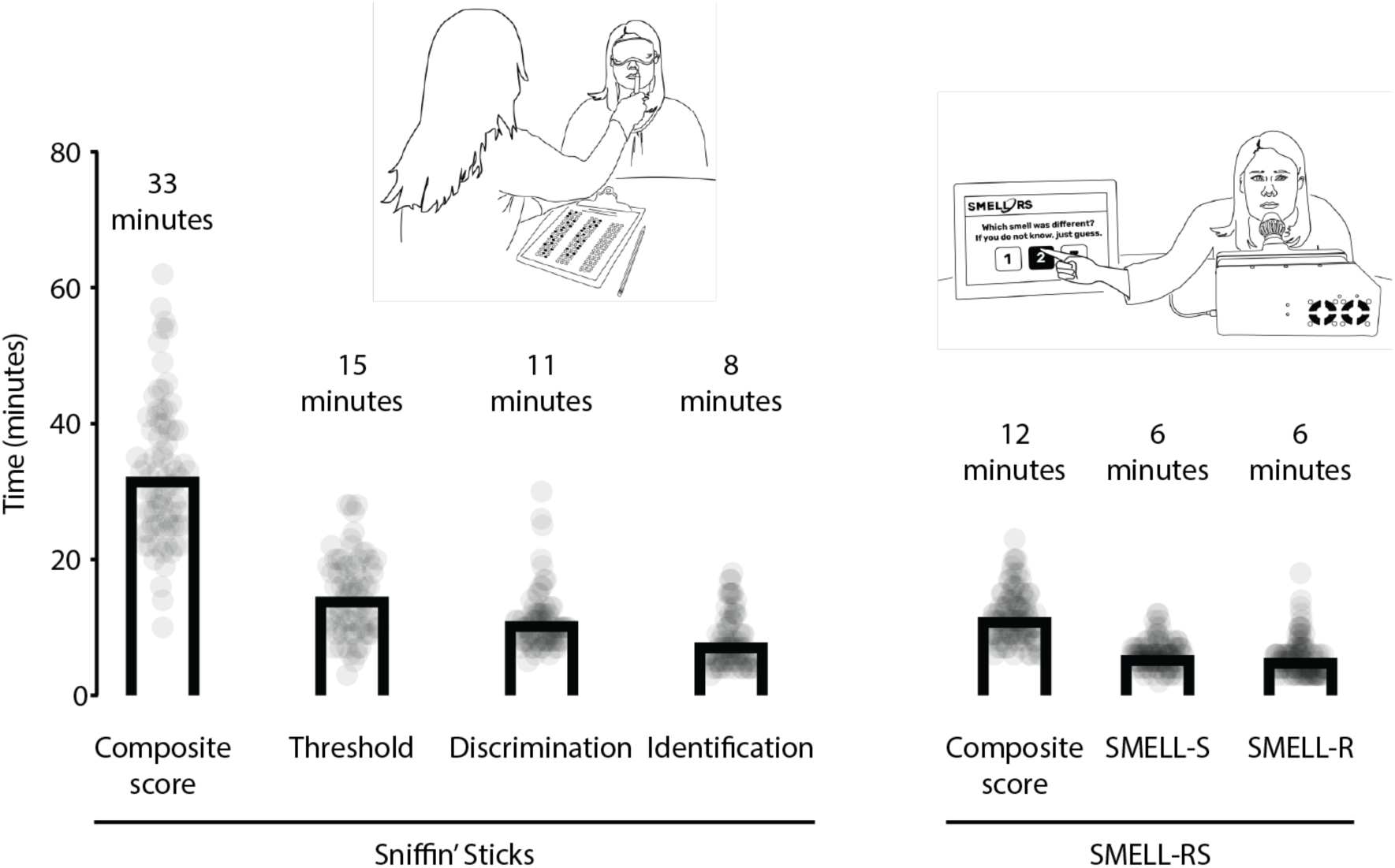
Test completion time. Each dot represents one subject, and the bars indicate the mean test completion time. Illustrations: M. Dougherty.

## Discussion

The present study demonstrates that the self-administered, digital version of SMELL-RS is (a) reliable and accurate; (b) potentially useful for differentiating the etiologies of olfactory loss; (c) capable of reducing misdiagnosis associated with phenylethyl alcohol Sniffin’ Sticks threshold testing; and (d) time-efficient, allowing evaluation of olfactory sensitivity (SMELL-S) or resolution (SMELL-R) in approximately six minutes per subtest without the need for trained personnel.

Evidence supporting the value of olfactory testing in daily rhinology practice continues to grow. Olfactory sensitivity has emerged as a clinically relevant marker of disease control in chronic rhinosinusitis (CRS)(22, 23). Preoperative assessment of olfactory function—and its responsiveness to corticosteroid trials—helps predict postoperative outcomes (24, 25). Knowledge of baseline olfactory function is also essential during functional endoscopic sinus surgery (FESS), particularly when addressing pathology involving the olfactory cleft (e.g., synechiae, narrow olfactory clefts, respiratory epithelial adenomatoid hamartomas, or polyposis)(26). When optimal surgical management and topical medical therapy fail to adequately control CRS, olfactory testing plays an important role in assessing indications for and response to emerging biologic therapies (27). Moreover, rhinologists are frequently confronted with non-CRS–related smell disorders, for which subjective self-ratings are well known to be unreliable (28). Despite this recognized need, formal olfactory sensitivity testing remains infrequently performed in routine practice. Our technology may solve this problem.

The self-administered, digital version of SMELL-RS demonstrated high test–retest reliability and showed a strong correlation with the Sniffin’ Sticks TDI composite score. Notably, SMELL-S alone performed similarly to the full SMELL-RS composite score, supporting its utility as a rapid, stand-alone test. Similar to how the pure-tone audiogram establishes hearing function, SMELL-S has potential to serve as a basic standardized olfactory assessment for routine ENT practice that is not only faster, but also more accurate than the gold standard threshold test. Interestingly, almost half of study subjects with the lowest score at the phenylethyl alcohol Sniffin’ Sticks threshold test had only moderately decreased or even normal olfactory function when measured with an odor-mixture based threshold test (SMELL-S) and other subjective or psychophysical measures. This may be due to variability in odor perception, a weaker smell of pure phenylethyl alcohol compared to pure SMELL-S mixture, and a more narrow phenylethyl alcohol serial dilution range that does not include concentrations that are high enough. We and others have previously shown that using odor-mixtures may lead to better test performance (9, 29). This finding has a large clinical impact because it uncovers the fact that a non-negligible proportion of patients may be misdiagnosed or may not benefit from an accurate follow-up testing after treatment.

SMELL-R evaluates a novel dimension of olfactory function by measuring odor resolution, defined as fine discrimination acuity across a graded difficulty scale. This performance domain is not adequately assessed by the traditional Sniffin’ Sticks Discrimination subtest, which is limited by ceiling effects and lacks a physical scale allowing systematic modulation of task difficulty. We hypothesize that this approach may be particularly valuable in patients presenting with persistent smell complaints, despite normal TDI composite scores. In such cases, deficits in higher-order resolution capacity may remain undetected using conventional testing. Furthermore, because SMELL-R represents a challenging and cognitively demanding task, we observed that some healthy participants failed SMELL-R testing. Several factors may explain this unexpected finding, including mental fatigue during prolonged testing, unfamiliarity with the tested odors, olfactory habituation, nose placement in relation to the device, sniffing times, and methodological differences related to odor delivery (constant airflow presentation versus headspace sampling from glass vials). These factors warrant further investigation to optimize testing protocols. Despite this, SMELL-R may serve as a sensitive screening tool to rule out early olfactory dysfunction associated with Parkinson’s disease or Alzheimer’s disease.

In the latest position paper on olfactory dysfunction published in 2023, the working group established that the unmet needs in smell testing were a test that is “quick and easy to administer” (2). Based on survey responses from 454 international ENT doctors, the main barrier to smell testing is insufficient time (3). By using a digital olfactometer, allowing self-administration coupled with SMELL-RS, we unlocked cross-cultural, rapid and automated olfactory testing without compromising on reliability or accuracy, nor olfactory sensitivity measurement, crucial information for ENTs, that is usually difficult to assess in clinical settings. Furthermore, automated medical reports can be easily generated and transferred to electronic medical records. Such systems can also be configured to perform even faster screening in a shorter period of time. According to Whitcroft and colleagues, 75% of international ENT doctors would be willing to perform a 12-minute evaluation for both subtests. Furthermore, 100% would perform a test that lasts under 5 minutes, which is only possible if they use SMELL-S (3). Other digital smell delivery devices for smell testing are currently under development, including Digital Scent (30), MultiScent-20 odor identification test (31, 32), mobile digital olfactory testing system (33), Sniff-Nano (34), Sony NOS-DX1000 (35). Devices should avoid cross-contamination between channels over time, be portable, allow threshold testing, be combined with universal and scalable smell testing methods, and affordable.

## CONCLUDING REMARKS

The digital version of SMELL-RS provides a reliable, accurate, and time-efficient method for comprehensive olfactory assessment, integrating both sensitivity (SMELL-S) and resolution (SMELL-R) measures. It could enable routine implementation with reduced personnel burden, detailed smell assessment or rapid screening, and generation of automated reports. It addresses major barriers to smell testing in clinical practice.

## Data Availability

All data produced in the present study are available upon reasonable request to the authors

## ACKNOWLEDGEMENTS

We thank our research volunteers for their time and interest in the study and the staff of the Rhinology-Olfactology Unit (Dehab Merke and Sandra Hausmann Jimenez) for their invaluable support. This work was performed with the contribution of the HUG NeuroCentre, Geneva University Hospitals, Geneva, Switzerland (Karl Schaller, Giannina Rita Iannotti, Alexie Ray, Nathalie Isidor).

## AUTHORSHIP CONTRIBUTION

J.W.H designed the study, analyzed the data, designed the figures, and wrote the manuscript. M.D prepared the odor-mixtures, designed the figures, wrote and reviewed the manuscript. A.P and D.B contributed to the design of the study, carried out the protocol, and reviewed the manuscript. P.S designed the study and reviewed the manuscript. R.H, D.P, E.M, M.O developed the hardware, wrote and reviewed the manuscript. L.B.V, A.K. and B.L contributed to the design of the study, analyzed the data, designed the figures, wrote, and reviewed the manuscript.

